# Population heterogeneity is a critical factor of the kinetics of the COVID-19 epidemics

**DOI:** 10.1101/2020.06.25.20140442

**Authors:** Dalkhat M. Ediev

**Affiliations:** North-Caucasian State Academy (Russia); Lomonosov Moscow State University (Russia); International Institute for Applied Systems Analysis (Austria)

## Abstract

The novel coronavirus pandemic generates extensive attention in political and scholarly domains ^1–4^. Its potentially lasting prospects, economic and social consequences call for a better understanding of its nature. The widespread expectations of large portions of the population to be infected or vaccinated before containing the COVID-19 epidemics rely on assuming a homogeneous population. In reality, people differ in the propensity to catch the infection and spread it further. Here, we incorporate population heterogeneity into the Kermack-McKendrick SIR compartmental model ^5^ and show the cost of the pandemic may be much lower than usually assumed. We also indicate the crucial role of correctly planning lockdown interventions. We found that an efficient lockdown strategy may reduce the cost of the epidemic to as low as several percents in a heterogeneous population. That level is comparable to prevalences found in serological surveys ^6^. We expect that our study will be followed by more extensive data-driven research on epidemiological dynamics in heterogeneous populations.

## Introduction

Because of the novelty and urgency of the situation, epidemiological models inform decision-making ^1,7–13^ in addressing the COVID-19 pandemic. Those models indicate high contagiousness of the virus and raise concerns about the majority of the population to be infected (if not vaccinated). The basic reproduction number *R*_0_ of the pandemic at its beginning was estimated to be around 3 ^9,14–16^, which implies 1−1/*R*_0_, i.e., about 67 percent of the population must be infected or vaccinated before the infection may be controlled without lockdown measures. This conclusion has affected mitigation policies in many countries, it has also contributed to expectations of recurrent waves of the epidemic.

Those models, however, ignore varying social engagement, epidemic-awareness, and hygiene preparedness that, along with other factors, contribute to the varying propensity of contracting the disease and spreading it to others. Various reports suggest 10-20 percent of cases may be responsible for 80 percent of the COVID-19 transmissions ^17–19^. These findings illuminate the fact that while the majority of people may barely contribute to the spread of the epidemics thanks to either limited social engagement or higher alertness and better hygiene, few others may become superspreaders infecting dozens of people. An essential practical conclusion from this conclusion was a call to aim the mitigation policies at superspreaders to reduce the basic reproduction number (average number of secondary infections per one initial infected person) and contain the spread of the infection. Another aspect of the heterogeneity, however, may demand to revise that conclusion and readdress the prospects of the pandemic and mitigation policies. The population heterogeneity is an essential player in the kinetics of the epidemic, because when the minority who contributes most to the spread of the virus contracts the disease and develops immunity, the outbreak may abruptly come to an end before the expected majority gets affected. Differential contagiousness also matters for how to manage the lockdown policies and whether to assume the recurrent waves of the epidemic after the lift of the social isolation measures or Autumn cooling. Furthermore, population heterogeneity may shorten the course of the outbreaks, because those with higher social engagement will also be the first to catch the infection.

## Results

In Figure 1, we present modeling results for three populations: the homogeneous population (the orange color), the heterogeneous population where 20 percent of those infected spread the virus in 80 percent cases (the blue color), and the heterogeneous population where 10 percent of infected are responsible for 80 percent of the secondary infections (the green color). Details of the simulation model are provided in the Supplementary Information. In all cases, the basic reproduction number is *R*_0_ = 3.

**Fig. 1.**
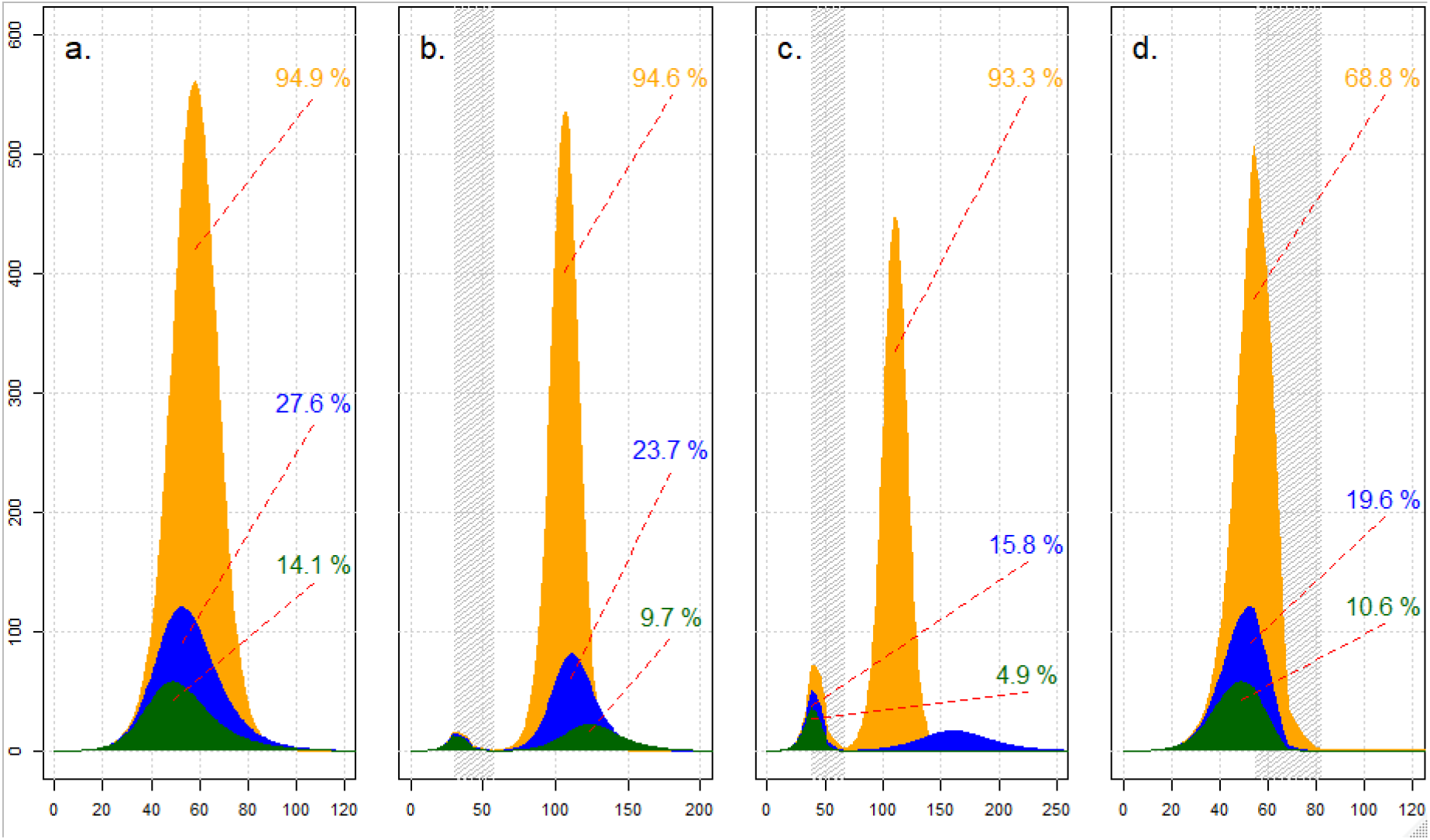
Evolution of the epidemics in three population scenarios: the homogeneous population (the orange color), the population where 20 percent of infected spread the virus in 80 percent cases (the blue color), and the population where 10 percent infected spread the infection in 80 percent cases (the green color). In all cases, the basic reproduction number is three. Horizontal axis: time (days) after 0.01 percent of the population gets infected; vertical axis: infected people per 1000 population; percents shown in the charts indicate proportions of the population infected throughout the epidemics. Pane **a**: no lockdown; panes **b**-**d**: lockdowns start in days 30, 44, and 55, respectively, last over 28 days, and reduce the spread of the infection by 90 percent (shaded areas indicate the lockdown periods).

In pane **a**, we present results for the evolution of the infected population (per 1000 original population) in scenarios with no lockdown and show numbers of ever-infected throughout the epidemic. With no policy intervention, the epidemic in the homogeneous population ends up with almost everybody (95 percent) infected. Under the basic reproduction number of three, the epidemics could have been checked after 67 percent of the population gets infected. In reality, that threshold may be surpassed thanks to the gained momentum of the spread of the infection. In the heterogeneous cases, the total numbers infected are also substantial (27.6 and 14.1 percent, respectively) but much lower than in the homogeneous case. The peak levels of the infected population are also much higher (56.1 percent) in the homogeneous population than in the heterogeneous ones (12.1 and 5.8 percent). Also note that the more heterogeneous is the population, the earlier is the peak of the epidemic. An intuition to this observation is that the faster infection (and recovery) of the superspreaders accelerates the epidemics in its early phase while slowing it down in a later phase.

In panes **b**-**d**, we present results for three timing options for the lockdown that lasts over 28 days and reduces the spread of the virus by 90 percent. When started too early (day 30, pane **b**), the lockdown leaves too many people susceptible to the virus and facilitates a substantial second wave. The total infected population is nearly the same as in the no-lockdown variant for the homogeneous population but considerably lower for the heterogeneous populations.

With a better timing of the lockdown, the long-term costs of the epidemics are much lower. The lockdown presented in pane **c** (starts in day 39) is optimal for the more heterogeneous population that experiences, with the optimal lockdown timing, no second wave (and the total number infected is minimal at 4.9 percent). That lockdown, however, is yet too early for the less heterogeneous population where a moderate second epidemic wave develops and leads to a total of 15.8 percent infected (a substantially higher cost as compared to the minimal cost of 11 percent associated with the lockdown starting in day 44). In the homogeneous case, the second wave is even higher, and almost everybody (93.3 percent) is, again, gets infected. Only a later lockdown that starts in day 55 (pane **d**) produces the optimal result for the homogeneous population (68.8 percent infected). While the first wave of the epidemics in the heterogeneous cases is earlier and more compressed as compared to the homogeneous case, the second wave, on the contrary, is later and more stretched out. Even if beneficial in terms of a lower peak, an extended small second wave may misguide the policymaker about the long-term efficiency of the lockdown measures.

Heterogeneous scenarios show much lower long-term costs of the epidemics and peak levels of the infected as compared to the traditional homogeneous case. If the lockdown had been more selective, better protecting the non-spreading population, those numbers could have been even smaller. Indeed, the epidemics could, in principle, be contained after most of the superspreaders were infected (bringing total infected populations down to about 1.5 and 0.3 percent in the two heterogeneous populations). On the other hand, a lockdown may also increase the cost of the epidemic if it selectively protects the superspreader population and results in many unnecessary infections among the non-spreaders.

Our modeling suggests that the optimally scheduled lockdown should only start at the moment when the susceptible population falls to such a level that the instantaneous reproduction number drops below the unity threshold. (See the Supplementary Information for the derivation of the reproduction number for the heterogeneous case.) After reaching that threshold, one should implement the strictest possible lockdown to cool off the epidemics’ momentum. That moment is earlier in the heterogeneous case for two reasons. Firstly, thanks to the higher infection rate and faster removal of the superspreaders from the susceptible population. Secondly, because of the faster decline of the reproduction number due to the compositional change that may only occur in a heterogeneous population. Milder but more prolonged lockdowns appear to be less efficient because longer-lasting epidemics result in more avoidable infections of the non-spreaders (simulations not shown here).

## Discussion

Expectations that about 70 percent of people may be infected before containing the pandemic were implicitly based on assuming population homogeneity. Contrary to those expectations, we show that the population heterogeneity may bring that threshold level down to as few as 14 percent with a similar basic reproduction number. Population heterogeneity, it appears, may even outweigh the vaccination in its importance as a factor checking the spread of the disease. We urgently need to fully understand the extent and nature of how people differ in susceptibility to the infection and the ability to spread it and appreciate that in our decision-making.

In the long run, a lower number of people infected means fewer causalities to the virus. In the short run, however, lockdown policies around the world take the capacity of the healthcare system into account too. In that context, it is notable that population heterogeneity also reduces the peak levels of the infected population (from 56 percent, as in the homogeneous case, to 6-12 percent).

Lockdown, when well-scheduled, is capable of substantially reducing the cost of the outbreak. The timing of the lockdown is crucial in all scenarios. When prematurely implemented, the lockdown leaves too large a portion of the population susceptible to the infection, which results in the second wave of the epidemic. In such cases, the epidemic may gain momentum and eventually lead to nearly the same total number of infected persons as in the case of no lockdown. The second wave appears to stretch over a more extended period in the heterogeneous cases, which may misguide policymakers in their assessment of the efficiency of the lockdown. Too late a lockdown, however, is also inefficient, because it allows for many avoidable infections.

In the optimal lockdown strategy, one should wait until the proportions susceptible fall to levels where the instantaneous reproduction number turns unity. After reaching that threshold, the lockdown measures should be implemented with maximal possible strength to cool off the epidemic’ momentum and halt the further spread of the virus. To design such an optimal policy response, however, it is mandatory to understand the kinetics of the epidemic well and assess the threshold correctly. That includes accounting for the role of population heterogeneity.

SIR-type epidemiological models and their extensions play an important role in designing policy responses to the COVID-19 and other epidemics. Our results imply that we should further extend those models to include different predispositions to catch and spread the infection.

With optimal lockdown strategy, the total number of infected people may be reduced to as low as five percent in the heterogeneous population. Notably, such level of prevalence is of the same magnitude as was found in serological surveys ^6^. Without undermining the importance of vaccine research, our results indicate the possibility of containing the COVID-19 pandemic without large-scale vaccinations.

Population heterogeneity also brings up the topic of selective lockdown. Both extreme selective lockdowns, the one protecting the non-spreading part of the population primarily and another one affecting only the superspreaders, may be realistic. On the one hand, there are all reasons to expect that those who are socially more engaged and less prepared to prevent the infection (such as epidemic deniers or hygienically less careful) will also be less affected by lockdown measures. That may contribute to the selectivity of the first type and reduction of the eventual cost of the epidemic. On the other hand, closing down public workplaces, introducing strict social distancing, isolation, or public hygiene measures may affect more the superspreaders while having limited effect, if at all, on socially less engaged and initially more prepared people. That may form lockdown selectivity of the second type and exaggerate the cost of the epidemic. Which scenarios develop in reality needs urgently being examined while countries move into the post-lockdown phases.

Long-term effects of the population heterogeneity reported here also call for revisiting the policy recommendations with respect to the superspreaders. The usual policy recommendation with respect to the superspreaders is to maximize lockdown efficiency among the superspreaders. Yet, we indicate that such a policy may delay but not prevent the second wave of the epidemic and spread, unnecessarily, the infection more into the non-spreaders population. We need to address this issue in designing social isolation policies.

The extent and type of population heterogeneity depend on many factors that need to be studied. Those include demographic factors such as age and sex, kinship structures and relations, household sizes and roles within them. Factors of heterogeneity also include biological predispositions, behavioral patterns (that, in turn, may depend on demographic circumstances, such as the presence of persons vulnerable to the disease in the household or kinship networks), educational, occupational, and income differentials, and others. A better understanding of these relations is instrumental in combating both the current urgency and other communicable diseases. To address those issues, however, we need representative and comparable statistics on how we differ in odds to catch and then to spread the virus. Such data are barely available, and we call statistical and healthcare agencies to urgently fill the gap in data on population heterogeneity.

## Data Availability

The results presented are based on modeling and simulations. The relevant R code is provided in the Materials and Methods.

## Supplementary. Materials and Methods

### The heterogeneous SIR model

We incorporate population heterogeneity into the discrete version of the Kermack-McKendrick SIR compartmental model ^5^. To distinguish contributions to the spread of the epidemic, we consider a set *K* of possible levels of the parameter of communicability *k* ∈ *K*. The latter is interpreted here as the number of people to whom an infected person passes the virus on at the beginning of the epidemic when nearly the entire population is yet susceptible to the infection. We term *k* ∈ *K* the communicability parameter (type *k* for brevity), although it may also reflect other factors that contribute to varying propensities of contracting and spreading the disease. Those factors may include varying levels of hygiene preparedness, behavioral patterns, occupation, kinship relations, household composition, etc. We disaggregate the population into groups of different levels of the communicability parameter:

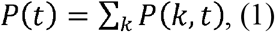

here,*P*(*t*) is the population size at time *tP*(*k, t*) is the size of the subpopulation of type k at time t; the summation is performed over all possible values of the communicability parameter *k* ∈ *K*. To model the spread of the infection, we consider the SIR model where the population consists of susceptible (*S*), infected (*I*), and recovered (*R*) compartments:

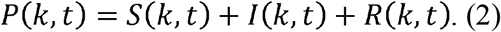

We assume a symmetric model where both the propensity of catching the virus and the propensity of spreading it are proportional to the communicability parameter *k*. Hence, we model new infections as follows:

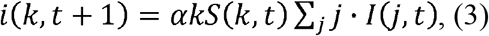

here,*i*(*k, t* + 1) is the number of new infections of persons of type *k* during the period *t*+ 1,*I*(*k, t*) is the stock of infected people of type *k* by the end of the period *t*; *α* is the model parameter that will further be related to the original susceptible population and the communicability period. The last multiplier in (3) is the communicability-weighted infected population that is not type-specific and may be abbreviated as *J* (*t*) = *∑*_*j*_*j*.*I*(*j, t*). Hence, *i*(*k, t* + 1) = *α kS*(*k, t*)*J*(*t*).

In modeling the course of recovering, we trace the duration of the infection period for the infected people and assume every infected person to recover in time *τ* after getting infected. That is, the number of people recovering in the period *t* equals:

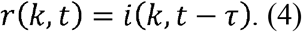

For simplicity, we assume that the communicability period is also equal to *τ*.

We conclude the model by applying balance equations to the stocks of susceptible, infected, and recovered populations:

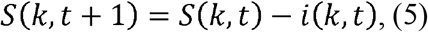

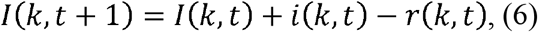

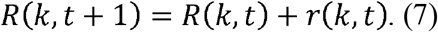

We neglect R-S transitions from recovered to the susceptible population because such transitions have not yet been reported to play a substantial role in the COVID-19 epidemics. We also neglect the fatality of the disease because we intend to highlight the primary effects of population heterogeneity upon the overall course of the epidemic. Introducing R-S transitions, mortality, and more realistic demographics should pose no difficulty in future research.

Assuming the entire original population is susceptible, the number of secondary infections per one initially infected person of type over the communicability period *τ* may be found from (3) as *τ α ∑*_*k*_ *kP*(*k*, 0)*j*. Given the interpretation of the communicability parameter, that number must be equal to, i.e., *τ α; ∑*_*k*_ *kP*(*k*, 0) = 1. That leads to the following estimate of the parameter *α*:

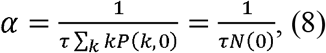

hereinafter, *N* (*t*) = *∑*_*k*_ *kS* (*k, t*) is the communicability-weighted susceptible population. The model of the infection’s spread may be rewritten as

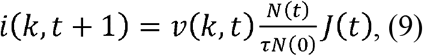

here, 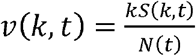 is the proportion of the weighted susceptible population of type k by the end of the period t. The total number of newly infected persons, 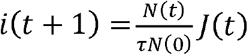, depends only on weighted susceptible and infected populations.

In the following, we model the effects of a lockdown by introducing a scaling coefficient *q* (*t*) that reflects the severity of social isolation, hygienic, and other mitigation measures in the period *t*:

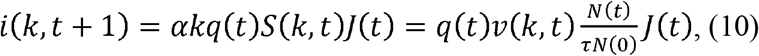

*q* = 1 indicates no efficient social isolation, while *q* = 0 corresponds to severe measures that halt the new infections completely. Eqs. (5) and (10) lead to the following closed-form solution for the evolution of the susceptible population of type *k*:

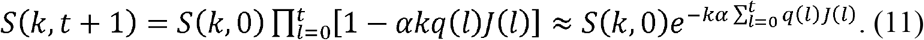

The higher the communicability parameter *k*, the faster is the fall of the susceptible population in (11). That creates compositional change in the remaining susceptible population, a change that suppresses the communicability-weighted susceptible population *J* (*t*) and checks the spread of the epidemics.

In generating and interpreting results of simulation scenarios, it is useful to relate the model parameters to the commonly used basic reproduction number *R*_0_. To establish the relation, assume the initial distribution of infected people follows the model relation (3) and is proportional to the weighted populations of each type: *i*(*j*, 0)∼*jP*(*j*, 0). The basic reproduction number, i.e., the secondary infections during period *τ* per one initial infection, equals:

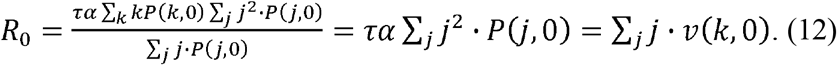

That is, the basic reproduction number is the weighted average of the communicability parameter with weights equal to the weighted susceptible populations of each type.

At an advanced phase of the epidemic, substantial portions of the population move to infected or recovered compartments, and the instantaneous reproduction number of a new outbreak decreases to:

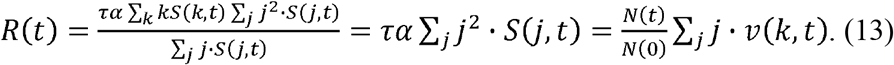

When *R* (*t*) ≤ 1, a new outbreak may be contained without a lockdown.

### Simulation details and additional simulations

We simulate epidemic scenarios under different assumptions about the population composition and the timing and the strength of a lockdown. The elementary time unit of simulations is one day. The communicability period *τ* = 14 days and the basic reproduction number *R*_0_ = 3 in all simulations. We assume either single or two subpopulations of different communicability parameter. In the latter (heterogeneous) case, the communicability parameter for the first subpopulation (‘non-spreaders’) is set at some sub-unity level *k*_1_ < 1; the parameter for the second (‘super-spreading’) subpopulation is set at such a level that the population-average basic reproduction number (12) equals 3. We form the heterogeneous populations in such a way that 10 or 20 percent of infected people are responsible for 80 percent of the further spread of the epidemic, similar to what was reported in the literature ^17–19^. Assuming that *x* percent of infected are responsible for *y* percent of transmissions, the communicability parameter of non-spreaders (*k*_1_) and superspreaders may be found from (12) as:

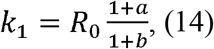

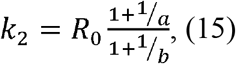

where 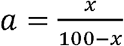 and 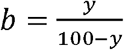 The proportion of the non-spreaders (*β*) is equal to:

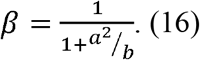

For the heterogeneous population, where 20 percent infected are responsible for 80 percent transmissions, *k*_1_ = 0.750, *k* = 12.1, and *β* = 0.985. For the more heterogeneous case where 10 percent of infected people transmit 80 percent of secondary infections, *k*_1_ = 0.667, *k* = 24, and *β* = 0.997.

Our simulations start with an initial 0.01 percent of the population infected. That initially infected population is distributed uniformly from 1 to *τ* in durations of contracting the disease. The initial distribution of the infected population by type is proportional to the weighted population of each type:*i*(*k*, 0)∼*kP*(*k*, 0). To assess the long-term costs of the epidemic, we run simulations over 40 communicability periods, i.e., 560 days.

We study two types of simulations: with and without a lockdown. In the scenarios of the main text, the lockdown lasts for 28 days, i.e. two communicability periods and reduces the spread of the infection by 90 percent (*q* = 0.1). In the following Figs. S1-S3, we present additional simulations with two options for the lockdown strength for each of the three populations considered: *q* = 0.1, i.e., reduction of the spread of the virus by 90 percent, and *q* = 0.001, i.e., almost a halt of the further spread of the virus. The duration of the lockdown in the figures (the shaded areas) is also 28 days.

First, consider the homogeneous population with the basic reproduction number R_0_ = 3, in which case our model turns to the conventional SIR model. In Fig. S1, we present sizes of the three population compartments in four selected simulations for the homogeneous population: no lockdown intervention (pane (a)); lockdown reducing the spread of the virus by 90 percent in the center the epidemic wave starting in period *t* = 45 and lasting for two communicability periods (pane (b), periods of lockdown are shaded in the figure); a similar lockdown that begins at time *t* = 56 found to be optimal in terms of reducing the numbers eventually infected (pane (c)); and the optimal lockdown of a similar timing as before but with a stronger effect (*q* = 0.001, pane (d)). Percentages of the total infected population throughout the entire simulation are indicated in each pane (*I*, percent of the initial population). In the absence of the lockdown, the model results in up to *I* = 95 percent of the homogeneous population getting infected by the end of the epidemic. That is considerably higher than the usually assumed proportion necessary to stop the spread of the disease (67 percent at *R*_0_ = 3). The latter proportion is surpassed thanks to the gained momentum of the epidemic when there is no lockdown or the timing of the lockdown is not optimal. Too early a lockdown leaves large portions of the population susceptible to the virus that fuels the second wave of the spread of the virus. The stricter the premature lockdown (simulation results not shown here), the later is the second wave, but the final proportion of people infected barely changes. The policy optimal for reducing the eventual number of infected people is a strict lockdown that starts when the proportion susceptible falls to 33 percent, the level sufficient to stop the further spread of the virus (pane (d)). Even a less strict lockdown with *q* = 0.1 that is similarly timed (pane (c)) leads to near-optimal results. Indeed, such an ‘optimality’ of the lockdown ignores infection fatality and healthcare systems’ capacity that has become a concern in many countries.

In Fig. S2, we present results for a scenario of the heterogeneous population where 20 percent of infected are responsible for 80 percent of transmissions. In this case, 1.5 percent of the population are superspreaders with the communicability parameter *k*_2_ = 12.1, while the majority of 98.5 percent are non-spreaders with *k*_1_ = 0.750. Even without a policy intervention (pane (a)), the spread of the epidemic stops at *I* = 27.6 percent of the population infected, less than half the level usually assumed for the basic reproduction number *R*_0_ = 3. Premature lockdown (staring at *t* = 40, pane (b)) is not sufficient to cool off the epidemic momentum completely (*I*= 14.7 percent), while optimally timed lockdown starting at *t* = 44 even if not strict enough (*q* = 0.1, pane (c)) results in almost an optimal outcome of *I*= 11.0 percent infected. The long-term cost of the epidemic is minimal (*I*= 9.7 percent) when the lockdown is both optimally timed, and strict enough (pane (d)).

An even more heterogeneous case where 10 percent of infected people spread the virus in 80 percent of the secondary cases is presented in Fig. S3. Here, only 0.3 percent of the population are superspreaders with *k* = 24.0; the rest are non-spreaders with *k*_1_ = 0.667. Without any policy intervention (pane (a)), the spread of the epidemic stops at only *I*= 14.1 percent of the population infected. The optimally timed strict lockdown (staring at *t* = 39, pane (d)) reduces the proportion of the infected people to as low as 4.2 percent. A weaker lockdown of similarly optimal timing is also efficient (*I* = 4.9 percent at *q* = 0.1, pane (c)). Premature lockdown (staring at *t* = 33, pane (b)), however, does not prevent the epidemic from gaining momentum and developing a stretched second wave (*I* = 8.0 percent, pane (c)).

Selected simulation results for other heterogeneous populations with *R*_0_ = 3 are presented in Table S1. Even with only half of the people being non-spreaders, and with no lockdown, the long-term cost of the epidemic and the peak number of the infected people decrease by more than 30 percent as compared to the homogeneous population. In an extreme case where 99.9 percent of people are non-spreaders, the long-term cost of the epidemic is only about five percent without any policy intervention. The peak level of the infected population also falls dramatically as the population heterogeneity increases.

**Table S1.**
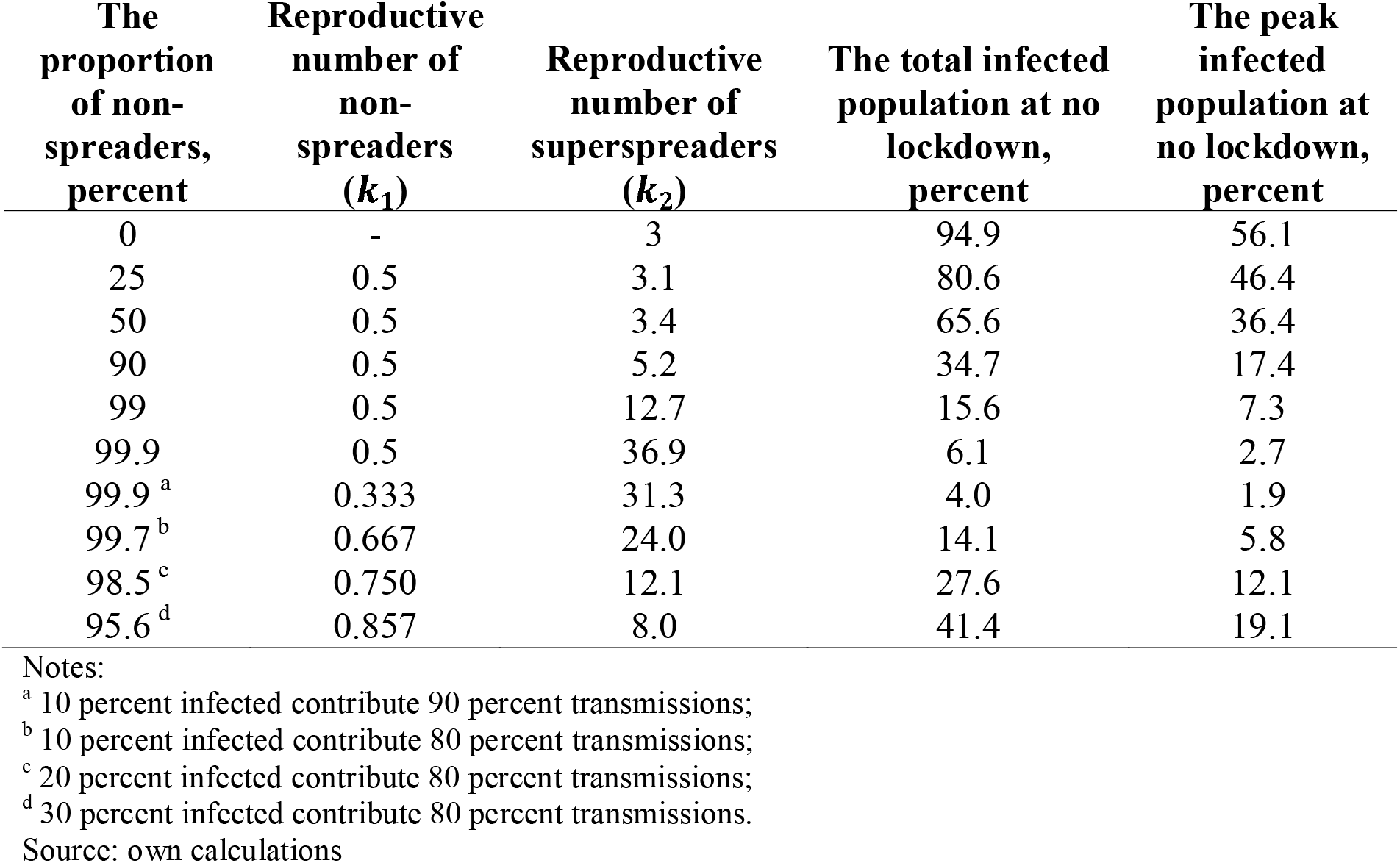
Selected simulation results for heterogeneous populations that consist of two subpopulations: non-spreaders with communicability parameter *k*_1_ < 1 and superspreaders with *k*_2_ > 1. In all cases, the basic reproduction number *R*_0_ = 3.

**Fig. S1.**
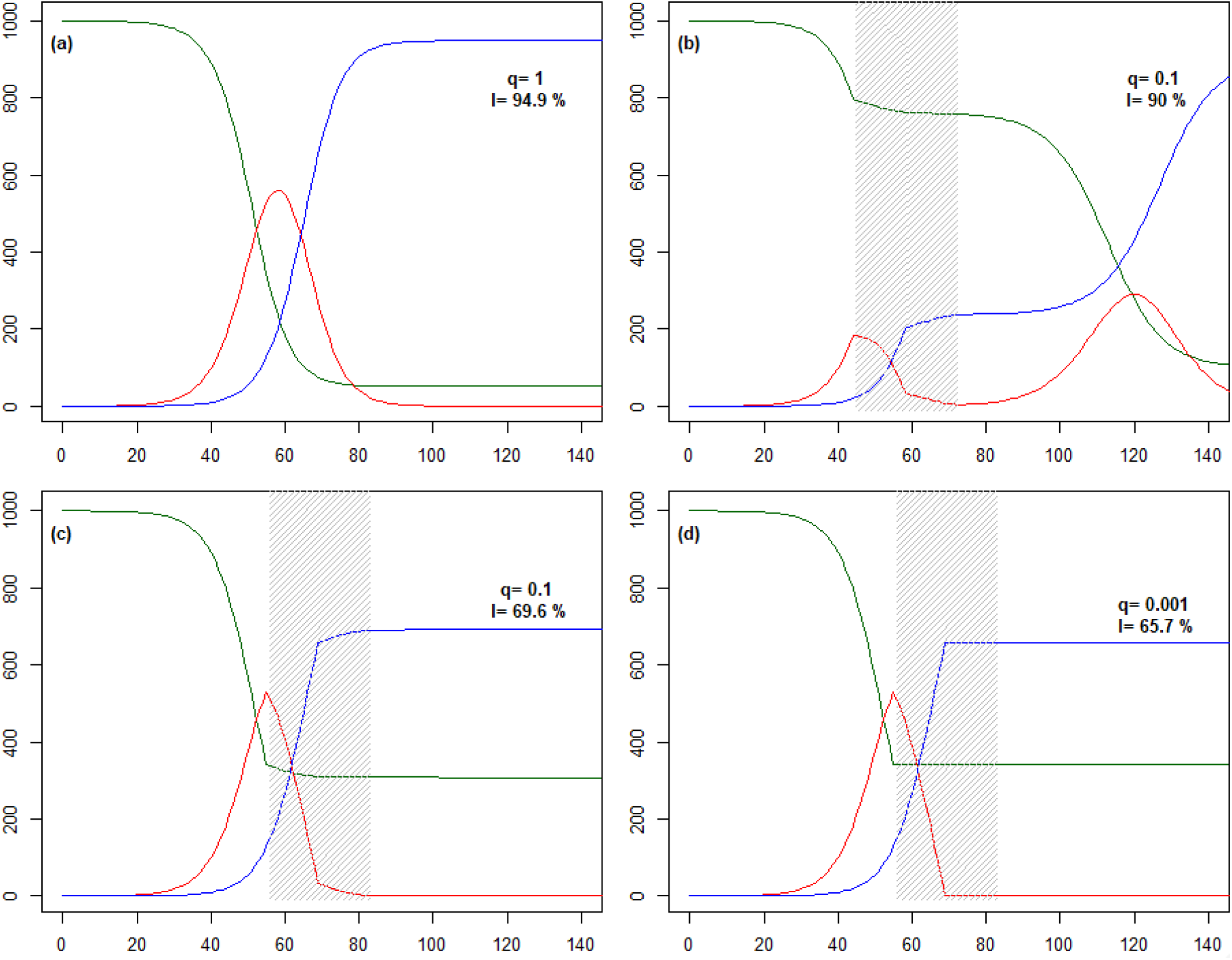
Simulation results for the homogeneous population with the basic reproduction number *R*_0_ = 3. Compartments susceptible (the green line), infected (the red line), and recovered (the blue line) populations per 1000 original population. Scenarios: no lockdown (pane (a)), lockdown reducing the spread by 90 percent that starts at *t* = 45 (pane (b)), lockdown reducing the spread by 90 percent that starts at *t* = 57 (pane (c)), and the optimal lockdown reducing the spread by 99.1 percent that starts at *t* = 57 (pane (d)). All lockdowns last for 14 days. Vertical axis: population size starting with 1000 original population. Horizontal axis: time in days from the original infection of 0.01 percent of people. *q* is the strength parameter of the lockdown; *I* is the eventual proportion of the population infected throughout the course of the epidemic.

**Fig. S2.**
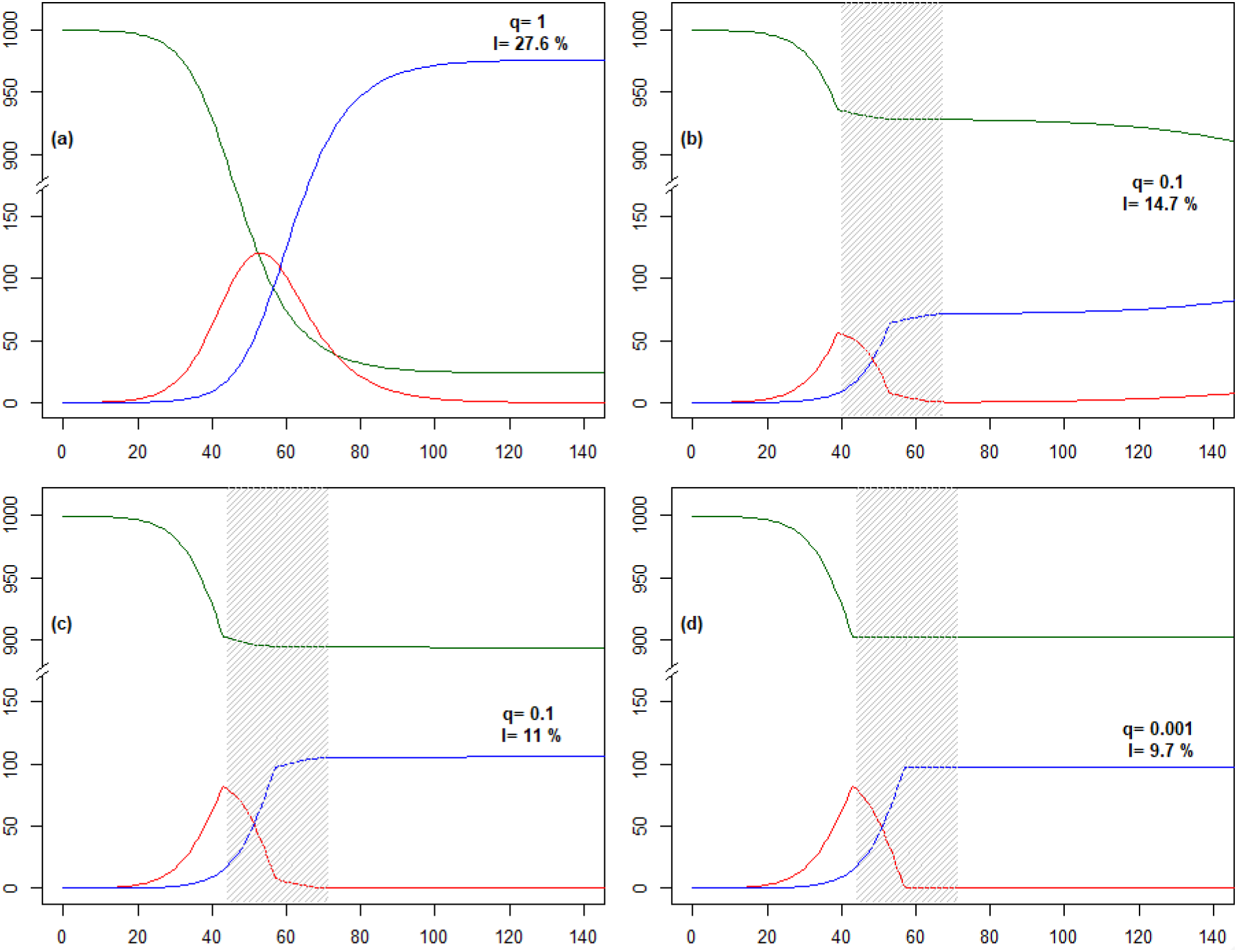
Simulation results for the heterogeneous population with the basic reproduction number *R*_0_ = 3 where 20 percent of infected people are responsible for 80 percent transmissions. 98.4 percent are non-spreaders with communicability parameter *k*_1_ = 0.750, and 1.6 percent are superspreaders with communicability parameter *k*_2_ = = 12.1. Compartments: susceptible (the green line), infected (the red line), and recovered (the blue line) populations per 1000 original population. Scenarios: no lockdown (pane (a)), a premature lockdown reducing the spread by 90 percent that starts at *t* = 40 (pane (b)), lockdown reducing the spread by 90 percent that starts at *t* = 44 (pane (c)), and the optimal lockdown reducing the spread by 99.1 percent that starts at *t* = 40 (pane (d)). All lockdowns last for 14 days. Vertical axis: population size starting with 1000 original population. Horizontal axis: time in days from the original infection of 0.01 percent of people. *q* is the strength parameter of the lockdown; *I* is the eventual proportion of the population infected throughout the course of the epidemic.

**Fig. S3.**
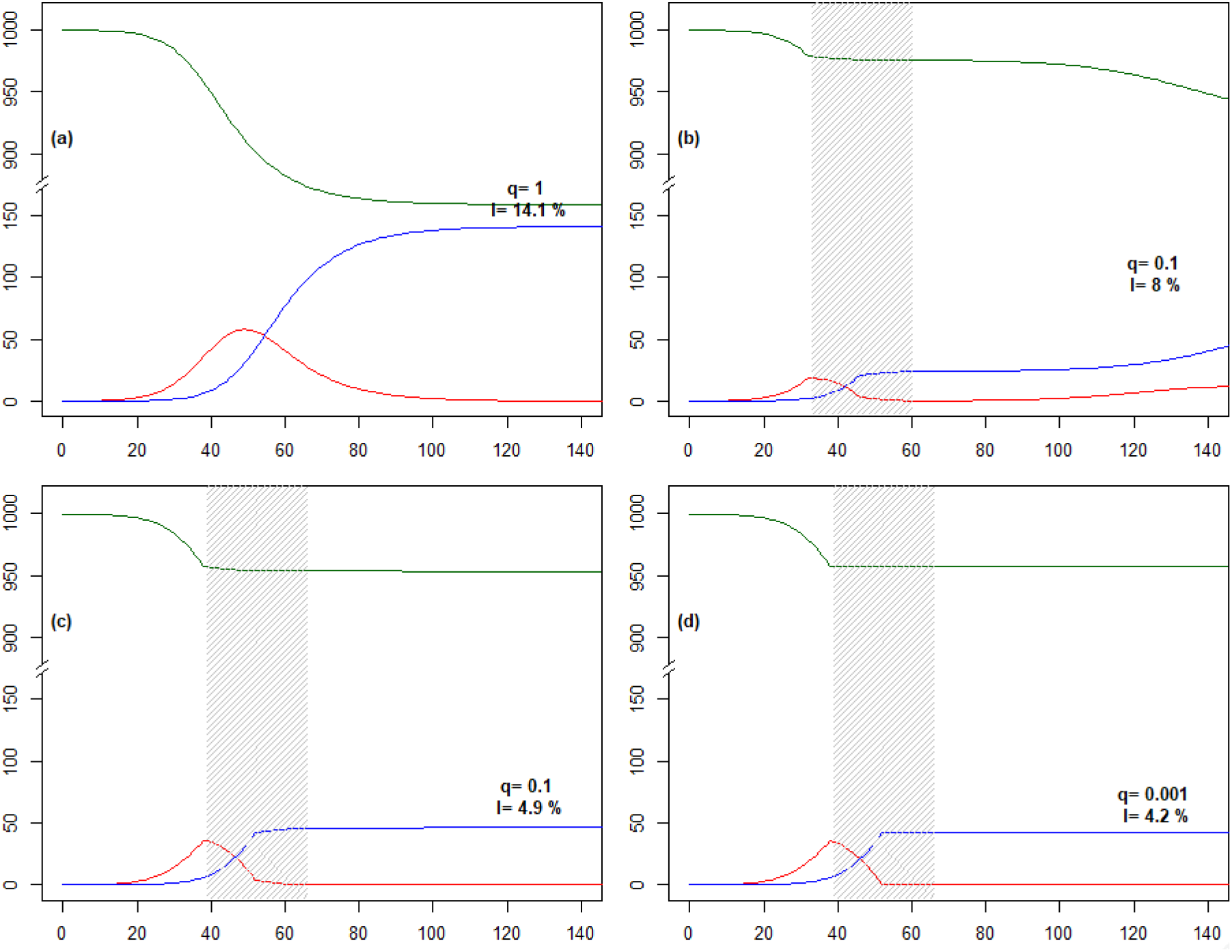
Simulation results for the heterogeneous population with the basic reproduction number *R*_0_ = 3 where 10 percent of infected people are responsible for 80 percent transmissions. 99.7 percent are non-spreaders with communicability parameter *k*_1_ = 0.667, and 0.3 percent are superspreaders with communicability parameter *k*_2_ = = 24. Compartments: susceptible (the green line), infected (the red line), and recovered (the blue line) populations per 1000 original population. Scenarios: no lockdown (pane (a)), a premature lockdown reducing the spread by 90 percent that starts at *t* = 33 (pane (b)), lockdown reducing the spread by 90 percent that starts at *t* = 39 (pane (c)), and the optimal lockdown reducing the spread by 99.1 percent that starts at *t* = 39 (pane (d)). All lockdowns last for 14 days. Vertical axis: population size starting with 1000 original population. Horizontal axis: time in days from the original infection of 0.01 percent of people. *q* is the strength parameter of the lockdown; *I* is the eventual proportion of the population infected throughout the course of the epidemic.

## R code to reproduce the simulations

~~~
#Celan code for the publication
P<-1000 #population size
R0<-3 #basic reproduction number
tau<-14 #communcability and recovery periods
step<-1 #modelling step, elementary time unit
Horizon<-1:(40*tau) #simulation horizon
i0<-0.0001 #proportion initially infected
I0<-i0*P #population initially infected
beta<-0.9 #propoirtion of non-spreaders; =0 for the homogeneous case
k1<-0.5 #communicability parameter for the non-spreaders
k2<-R0*(1+sqrt(1+4*beta*k1*(R0-k1)/(R0^2*(1-beta))))/2 #communicability parameter
for the superspreaders
K<-c(k1,k2) #communicability parameters
~~~

~~~
#ditribution of the population by the communicability parameter
distr<-rep(0,length(K))
distr[1]<-beta
distr[2]<-1-beta
#more types may be added if necessary
~~~

~~~
#lockdown parameters
Qt<-47 #beginning of lockdown
Quaranteen<-Qt+c(1:(2*tau))-1 #period of the lockdown
qq<-0.1 #lockdown efficiency coef. 1 - no lockdown; 0 - absolute halt of epidemic
Q<-ifelse(Horizon %in% Quaranteen,qq,1)
#generate scenario
Pkt<-CoVgen(P=P,K=K,distr=distr,I0=I0,Q=Q,Horizon=Horizon,tau=tau,step=step)
#summary and plot
Pt.agr<-aggregate(Pkt[,-1], by=list(t=Pkt$t), FUN=sum)
plot(Pt.agr$t,Pt.agr$S,type=“l”,col=“darkgreen”,
   ylim=c(0,P),xlim=c(0,20*tau/step),xlab=““,ylab=““)
if(is.na(qq)|qq<0) qq=“lim.prev.”
lines(Pt.agr$t,Pt.agr$I,col=“red”)
lines(Pt.agr$t,Pt.agr$R,col=“blue”)
R.final<-Pt.agr[Pt.agr$t==max(Horizon),]$R
~~~

~~~
if(min(Q)<1) rect(min(Horizon[Q!=1]),-10,max(Horizon[Q!=1]),
          1100,density=30,col=“grey”,border=NA)
text(x=9*tau/step,y=min(0.75*P,R.final),paste(“q=“,qq,”\nI=“,paste(round(100*R.final/P,1),”%”)),pos=3,font=2)
~~~

~~~
#function generating the epidemic scenario for the given set of parameters
CoVgen<-function(P,K,distr,I0,Q,Horizon=1:length(Q),tau=14,step=1) {
 istr<-distr/sum(distr)
 I.matr<-matrix(rep(0,length(K)*tau),ncol=tau)
 P.k<-P*distr
 N0<-sum(K*P.k)
 alpha<-step/(tau*N0)
 I.k<-pmin(P.k,I0*K*P.k/N0)
 if(sum(I.k)<I0) {
  P.resid<-P.k-I.k
  I.k<-I.k+(I0-sum(I.k))*P.resid/sum(P.resid)
 }
 I.matr[,1:tau]<-I.k/tau
 I.k<-rowSums(I.matr)
 I<-sum(I.k)
 J<-sum(K*I.k)
 S<-P.k-I.k
 N<-sum(K*S) R0.effective<-(J/I)*N/N0
 Pkt<-data.frame(t=0,k=K,P=P,S=S,I=I.k,R=0,
           N=N,J=J,R0=R0,
           R0.effective=R0.effective)
 for (t in Horizon) {
  q<-Q[Horizon==t] #lockdown parameter: the lockdown reduces alfa q-fold
  S.prev<-Pkt[Pkt$t==t-1,]$S
  R.prev<-Pkt[Pkt$t==t-1,]$R
  I.prev<-Pkt[Pkt$t==t-1,]$I
  J.prev<-sum(K*I.prev)
  I.k.new<-pmin(S.prev,q*alpha*K*S.prev*J.prev)
  I.rec<-I.matr[,tau]
  I.matr[,2:tau]<-I.matr[,1:(tau-1)]
  I.matr[,1]<-I.k.new
  I.k<-rowSums(I.matr)
  S<-S.prev-I.k.new
  R<-R.prev+I.rec
  I<-sum(I.k)
  J<-sum(K*I.k)
  N<-sum(K*S)
  R0.effective<-(J/I)*N/N0
  Pkt<-rbind(Pkt,
        data.frame(t=t,k=K,P=P,S=S,I=I.k,R=R,
               N=N,J=J,R0=R0,
               R0.effective=R0.effective))
 }
 return(Pkt)
}
~~~

## Notes

### Competing Interest Statement

The authors have declared no competing interest.

### Funding Statement

This research has been based on no external funding.

